# An estimation of global genetic prevalence of PLA2G6-associated neurodegeneration

**DOI:** 10.1101/2023.12.21.23300352

**Authors:** Amina Kurtovic-Kozaric, Moriel Singer-Berk, Jordan Wood, Emily Evangelista, Leena Panwala, Amanda Hope, Stefanie M. Heinrich, Samantha Baxter, Mark J. Kiel

**Affiliations:** Genomenon, Inc., 206 E. Huron St. Suite 114, Ann Arbor, MI 48109, USA; Broad Institute of MIT and Harvard, Cambridge, MA 02141, USA; The INADcure Foundation, Fairfield, NJ 07004, USA

**Keywords:** PLA2G6-associated neurodegeneration, INAD, genetic prevalence

## Abstract

**Background:** PLA2G6-associated neurodegeneration (PLAN) comprises three diseases with overlapping features: infantile neuroaxonal dystrophy (INAD), atypical neuroaxonal dystrophy (atypical NAD), and PLA2G6-related dystonia-parkinsonism. INAD is an early onset disease characterized by progressive loss of vision, muscular control, and mental skills. The prevalence of PLA2G6-associated diseases has not been previously calculated.

**Methods:** To provide the most accurate prevalence estimate, we utilized two independent approaches: database-based approach which included collecting variants from ClinVar, Human Gene Mutation Database (HGMD) and high confidence predicted loss-of-function (pLoF) from gnomAD (Rare Genomes Project Genetic Prevalence Estimator; GeniE), and literature-based approach which gathered variants through Mastermind Genomic Search Engine (Genomenon, Inc). Genetic prevalence of PLAN was calculated based on allele frequencies from gnomAD, assuming Hardy–Weinberg equilibrium.

**Results:** In the PLA2G6 gene, our analysis found 122 pathogenic, 82 VUS, and 15 variants with conflicting interpretations (pathogenic vs VUS) between two approaches. Allele frequency was available for 58 pathogenic, 42 VUS, and 15 conflicting variants in gnomAD database. If pathogenic and VUS variants are included, the overall genetic prevalence was estimated to be 1 in 220,322 pregnancies, with the highest genetic prevalence in African/African-American populations at 1 in 86,012 pregnancies. Similarly, the highest carrier frequencies observed were in African/African-American and Asian populations.

**Conclusion:** Our estimates highlight the significant underdiagnosis of PLA2G6-associated neurodegeneration and underscores the need for increased awareness and diagnostic efforts. Furthermore, our study revealed a higher carrier frequency of PLA2G6 variants in African and Asian populations, stressing the importance of expanded genetic sequencing in non-European populations to ensure accurate and comprehensive diagnosis. Future research should focus on confirming our findings and implementing expanded sequencing strategies to facilitate maximal and accurate diagnosis, particularly in non-European populations.

## Background

PLA2G6-Associated Neurodegeneration (PLAN) comprises three autosomal recessive diseases with overlapping features: infantile neuroaxonal dystrophy (INAD, OMIM #256600, ORPHA: 35069), atypical neuroaxonal dystrophy (atypical NAD, OMIM #610217)), and PLA2G6-related dystonia-parkinsonism (PARK14, OMIM #612953), which are caused by pathogenic variants in the PLA2G6 gene (1-4). PLA2G6 encodes an enzyme called phospholipase A2 group VI, which plays a vital role in lipid metabolism and maintaining the integrity of cell membranes, especially in nerve cells (2). Pathogenic variants in the *PLA2G6* gene lead to the accumulation of abnormal lipids in neuronal tissues. This lipid accumulation, particularly in axons, can cause structural damage and impair the transmission of nerve signals.

INAD is an early onset disorder and is associated with severe symptoms including ataxia, mental and motor deterioration, hypotonia, progressive spastic tetraparesis, visual impairments, bulbar dysfunction, and extrapyramidal signs (4). These symptoms are progressive and INAD patients usually succumb to the disease before their 10th birthday. Atypical cases, such as aNAD and PARK14, have a later onset of symptoms, including progressive dystonia and parkinsonism. Cerebellar atrophy is a characteristic symptom in both INAD and aNAD (5,6).

Iron accumulation in the basal ganglia has been observed in some INAD, aNAD, and PARK14 patients, leading to the classification of these diseases as neurodegeneration with brain iron accumulation 2 (NBIA2) (7). The neuropathological hallmarks of PLAN include the formation of spheroid structures containing membranes, α-synuclein, and ubiquitin, known as tubulovesicular structures (TVSs) (8). Additionally, Lewy bodies and phosphorylated tau-positive neurofibrillary tangles have been observed in the nervous system of PLAN patients (9).

The diagnosis of INAD includes a combination of clinical evaluation, neurological examination, electrophysiology, imaging tests, as well as skin biopsies revealing axonal swellings and spheroid bodies in pre-synaptic terminals within the central or peripheral nervous system (10,11). Confirmatory biopsies often require multiple attempts, resulting in prolonged waiting periods for families seeking a diagnosis. Advancements in genetic testing paired with decreasing cost of gene and genome sequencing have expedited the diagnostic process. Families now receive diagnoses more quickly, sometimes within a year of initial symptom onset (12), but challenges remain. Although genome sequencing (GS) has the capacity to identify large and complex variants, interpreting these variants continues to pose difficulties, particularly when dealing with non-coding variants (13). Challenges associated with identifying the causative variants from exome sequencing (ES)/GS can be broadly categorized into two groups: 1) interpretation issues, such as variants of uncertain significance (VUS) in known or novel disease genes and non-coding variants, and 2) detection challenges, including the absence of a second variant in recessive disorders or the presence of causative variants in regions that are difficult to sequence or are masked due to biases in reference genomes and genomic datasets. Some very rare variants may be unobserved in large population databases and do not have associated MAF (minor allele frequency) (14).

These challenges in variant identification and interpretation affect the diagnosis and the estimation of prevalence of PLA2G6-related disorders, which is currently unknown, but ranges from ∼1 in 1,000,000 from the Rare Genomics Institute to 1 in 1-2 million children (15-18).

The goal of this study was to improve previous estimates by performing two independent approaches to identify pathogenic variants associated with PLA2G6-associated neurodegeneration: 1) an extensive literature review utilizing Mastermind Genomic Search Engine (19) to collect additional variants and supporting evidence, and 2) extracting data from publicly existing databases such as ClinVar, HGMD and gnomAD (18). For both approaches, we employ standard variant interpretation to evaluate the level of evidence supporting the inclusion of variants in the estimate of disease prevalence (20,21,22).

## Methods

### Study design and participants

To evaluate the genetic spectrum of the PLA2G6 variants, a comprehensive search was performed to identify all previously reported disease-causing *PLA2G6* gene variants using two independent approaches. One approach used Mastermind Genomic Search Engine (Genomenon, Inc.) (19) exclusively, while the second approach developed by the Rare Genomes Project’s Genetic Prevalence Study (15) combined several publicly available resources: gnomAD, HGMD, and ClinVar.

The inclusion criteria for targeted literature review conducted through Mastermind were: 1) studies published as full-text publications; 2) English or other languages were included; 3) single nucleotide variants and indels; 4) published until 30 April 2023. The terms included in the search strategy in Mastermind were defined by automated indexing using genomic language processing (18). Mastermind search for published variants in *PLA2G6*, including single-nucleotide variants (SNVs) and indels, yielded 379 articles. Once articles were collected, variants were extracted and duplicate variants removed. Benign variants were removed from further analysis (Figure 1). The remaining variants were curated and classified following current ACMG/AMP Guidelines for Sequence Variant Interpretation and ClinGen specifications (21,22).

**Figure 1.**
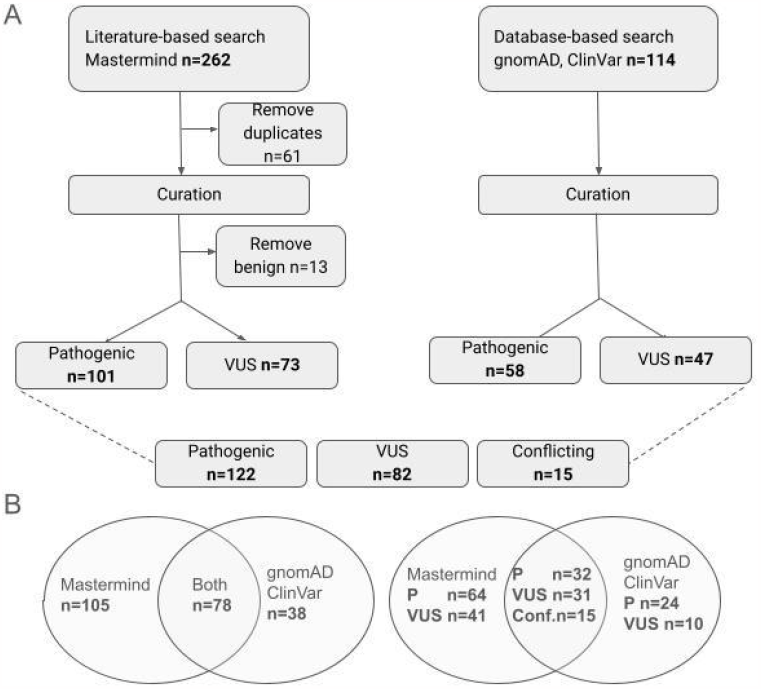
(A) To estimate genetic prevalence of PLA2G6-associated neurodegeneration, we have utilized a literature-based approach (left) and database-based approach (right). In total, 122 pathogenic, 82 VUS, and 15 conflicting variants were curated and classified. (B) Pathogenic and VUS variants recorded by two independent approaches.

The Rare Genomes Project’s Genetic Prevalence Study approach combined variants found in several databases: ClinVar (pathogenic, likely pathogenic and conflicting where one of the submissions listed the variant as pathogenic or likely pathogenic), HGMD (DM) and gnomAD high confidence (HC) predicted loss-of-function (pLoF). Benign (B) and likely benign (LB) variants were removed. Variants that were labeled as conflicting were removed if the conflict was LB vs L, VUS vs LB/B, while conflicting LP (likely pathogenic) vs P (pathogenic) vs VUS were kept for further classification following current ACMG/AMP Guidelines for Sequence Variant Interpretation and ClinGen specifications (20). Additionally, high confidence predicted loss-of-function (pLoF) variants in gnomAD, including those in the list above, underwent a pLoF curation (15). The pathogenic and likely pathogenic variants were further selected, as illustrated in Figure 1. The variants were extracted from gnomAD v2.1.1 along with their overall and population-specific allele frequencies (23). gnomAD v2.1.1 contains 125,748 exome sequences and 15,708 whole-genome sequences from 141,456 unrelated individuals. It is of note that gnomAD attempts to remove cohorts that were recruited for pediatric disease. Given the substantial morbidity associated with PLA2G6-associated neurodegeneration, we made the assumption that the gnomAD dataset does not include any individuals with this disease. If homozygotes were detected, they were removed during the curation process, as described below.

Each variant was standardized using the GRCh37/hg19 genome build and the canonical transcript— NM_003560.4—as well as the nomenclature guidelines set by the Human Genome Variation Society (HGVS) (21). As described in Figure 1, the curation process involved manually curating selected variants according to the standards set by the American College of Medical Genetics and Association of Molecular Pathologists (ACMG/AMP) into benign (B), likely benign (LB), variant of unknown significance (VUS), likely pathogenic (LP) and pathogenic (P) (22). This interpretation process considered clinical and functional studies from the literature, population frequencies derived from gnomAD v2.1.1, computational predictions of the effect of missense variants derived from REVEL, PolyPhen-2, MutationTaster2, and SIFT, and computational predictions of splicing defects for single nucleotide variants derived from dbscSNV (26-30). Once variants were identified and curated from two independent approaches, they were combined into a single database.

#### Allele frequency and prevalence calculations

Genetic prevalence estimates were produced according to a previously described method (24). The disease prevalence was estimated by using the observed allele frequency of a pathogenic/likely pathogenic variant in the gnomAD database as the direct estimator for qi, as described previously in Shourick et al (31). Genetic and disease prevalences were calculated separately for pathogenic, VUS, and conflicting variants. The overall (or population-specific) allele frequency was summed across all selected variants and then used within the Hardy–Weinberg equation to calculate the carrier frequency (2pq) and the frequency of a disease-causing genotype (q^2^) (26).

## Results

Two independent approaches were utilized to improve yield of pathogenic PLA2G6 variants to estimate genetic and disease prevalence of PLA2G6-associated neurodegeneration. Comprehensive retrieval of pathogenic variants using Mastermind resulted in the identification of 379 articles, which yielded 262 variants. As shown in Figure 1A, 61 variants were removed because they were duplicates. The remaining 201 variants were curated and classified according to ACMG/AMP guidelines (22), which resulted in removal of 13 benign variants. After the curation, 101 pathogenic variants and 73 VUS were identified and added to the database for further analysis. Using RGP’s Genetic Prevalence Study approach based on publicly available databases, 114 variants were identified (Figure 1A). The variants were curated according to ACMG/AMP guidelines and pLoF curation framework, yielding 58 likely pathogenic/ pathogenic variants and 47 VUS, which were added to the database of PLA2G6 variants. Once the curation process was completed, an additional step was taken to analyze the more common variants that were curated as VUS, but were rarely seen in the patient population or were associated with other similar diseases, such as p.Tyr319Met or p.Pro702Ser; these variants were removed from further analysis. Curated variants were combined into a single database of 122 pathogenic, 82 VUS, and 15 variants with conflicting interpretations between the approaches. This includes all variants, including those variants that have zero allele frequency in gnomAD (Supplementary Table 1). In total, 115 variants had allele frequency reported in gnomAD. The overlapping variants between two approaches are presented in Figure 1B.

Assessing the contribution of the most common pathogenic and VUS PLA2G6 variants to the genetic prevalence estimate with non-zero allele frequency in gnomAD, revealed that 15 variants accounted for 50% of the total allele frequency, namely p.Ala147Thr, p.Arg6Cys, p.Arg677Leu, p.Tyr790*, p.Arg741Gln, p.Arg301Cys, p.Arg70Gln, p.Val421AlafsTer26, p.Asp331Tyr, p.Leu129Pro, p.Arg710His, p.Arg635*, p.Arg635Gln, and p.Arg37*. The top five most frequent variants alone (p.Ala147Thr, p.Arg6Cys, p.Arg677Leu, p.Tyr790*, and p.Arg741Gln) accounted for 24% of all variants. If pathogenic variants with non-zero allele frequency are analyzed (n=58), 9 variants accounted for 50% of the total allele frequency, namely p.Tyr790* (c.2370T>G), p.Val421AlafsTer26, p.Arg635Ter, p.Arg635Gln, p.Arg600Gln, p.Arg37Ter, p.Tyr790* (c.2370_2371del), c.209+2T>G, and Gly638Arg (Figure 2B). Besides allele frequencies, cumulative percentage of genetic prevalence of VUS and pathogenic variants (Fig. 2C) and pathogenic variants only (Fig. 2D) with non-zero allele frequencies has also been presented to show the share of each variant in genetic prevalence of PLA2G6-associated neurodegeneration and neuroaxonal dystrophy.

**Figure 2A:**
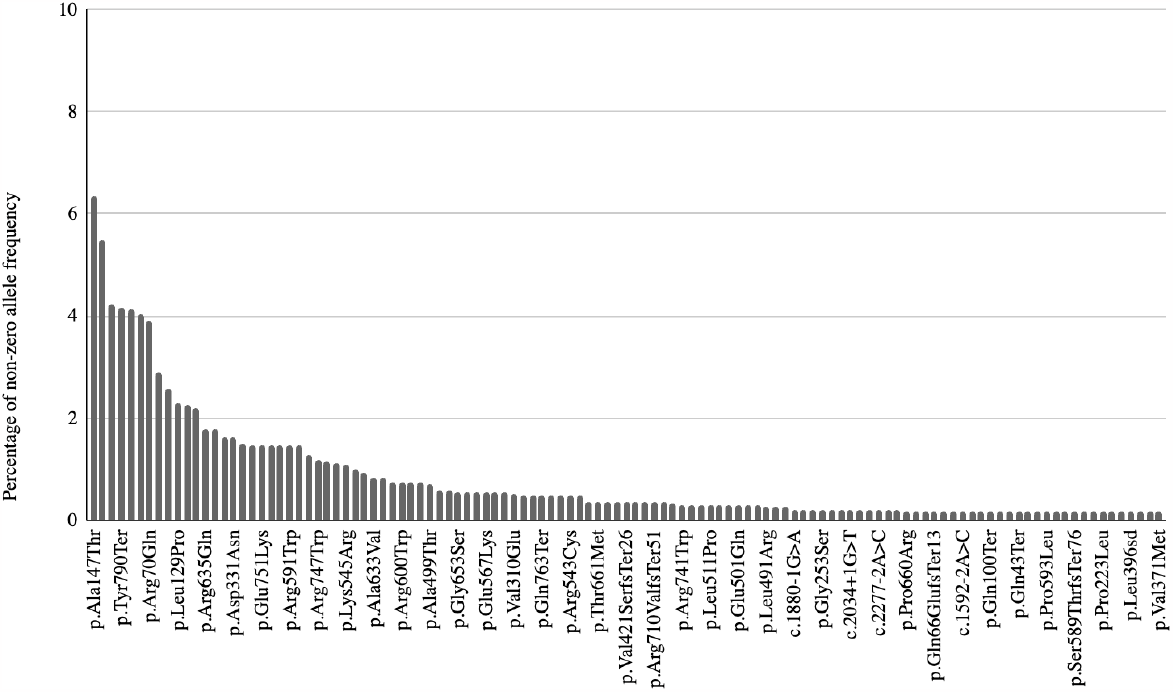
Percentage of allele frequency of all pathogenic and VUS variants in PLA2G6 with non-zero allele frequencies

**Figure 2B:**
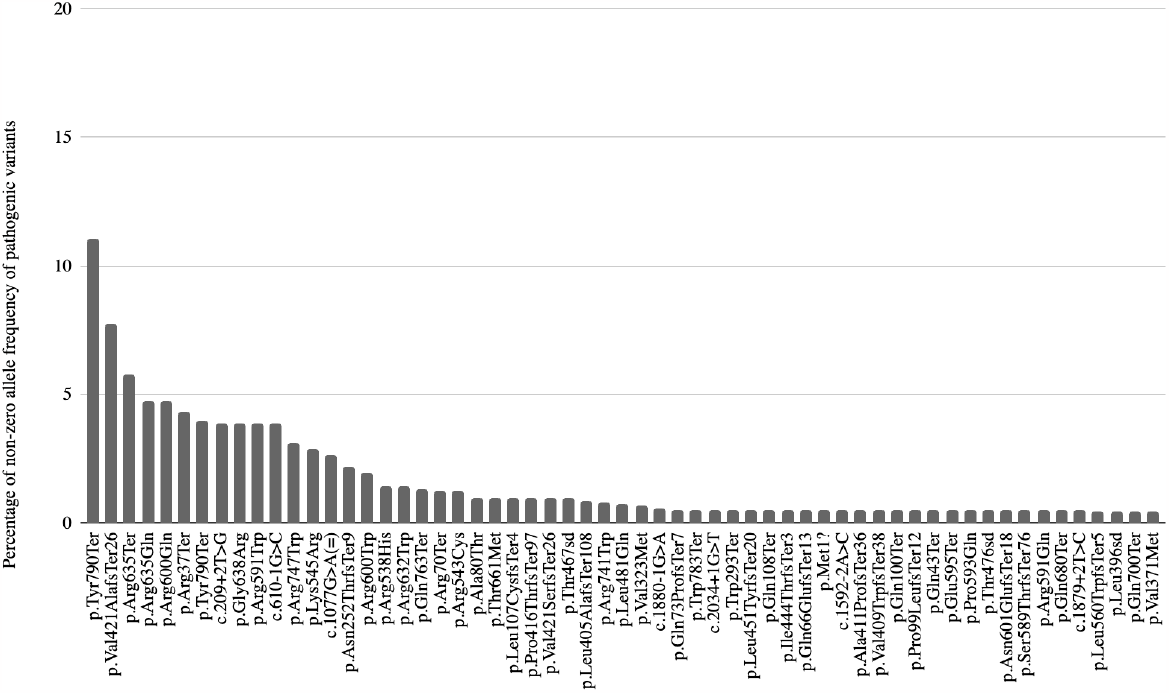
Percentage of allele frequency of all pathogenic variants in PLA2G6 with non-zero allele frequencies

**Figure 2C:**
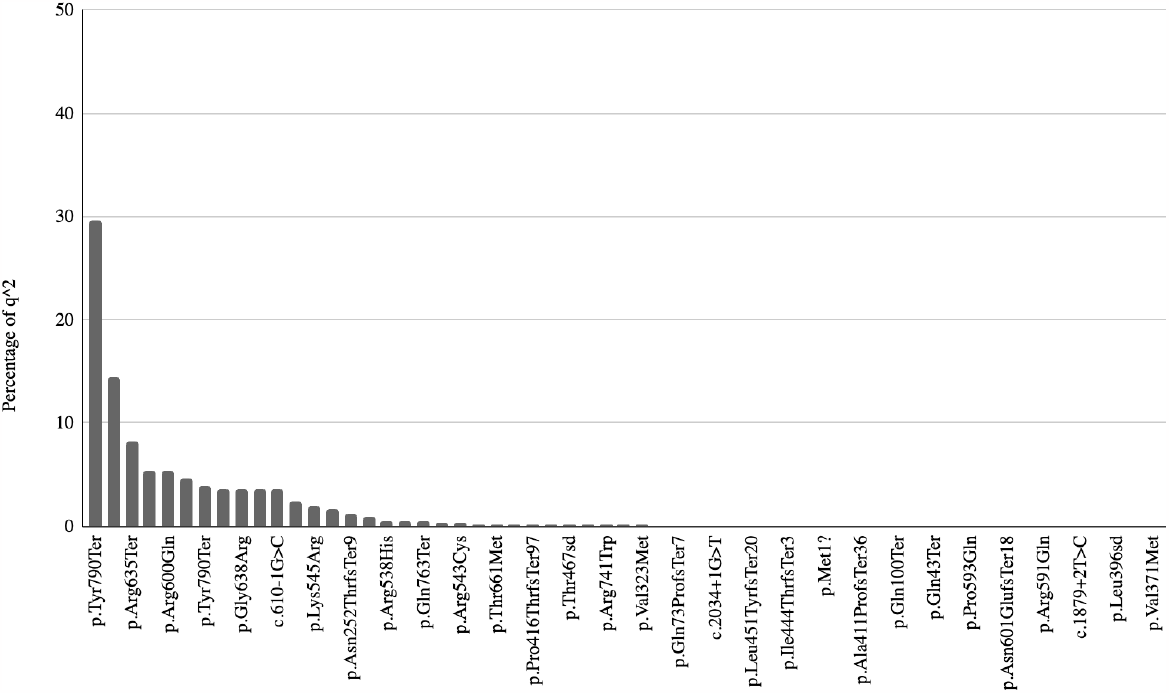
Percentage of genetic prevalence of VUS and pathogenic variants in PLA2G6 with non-zero allele frequencies

**Figure 2D:**
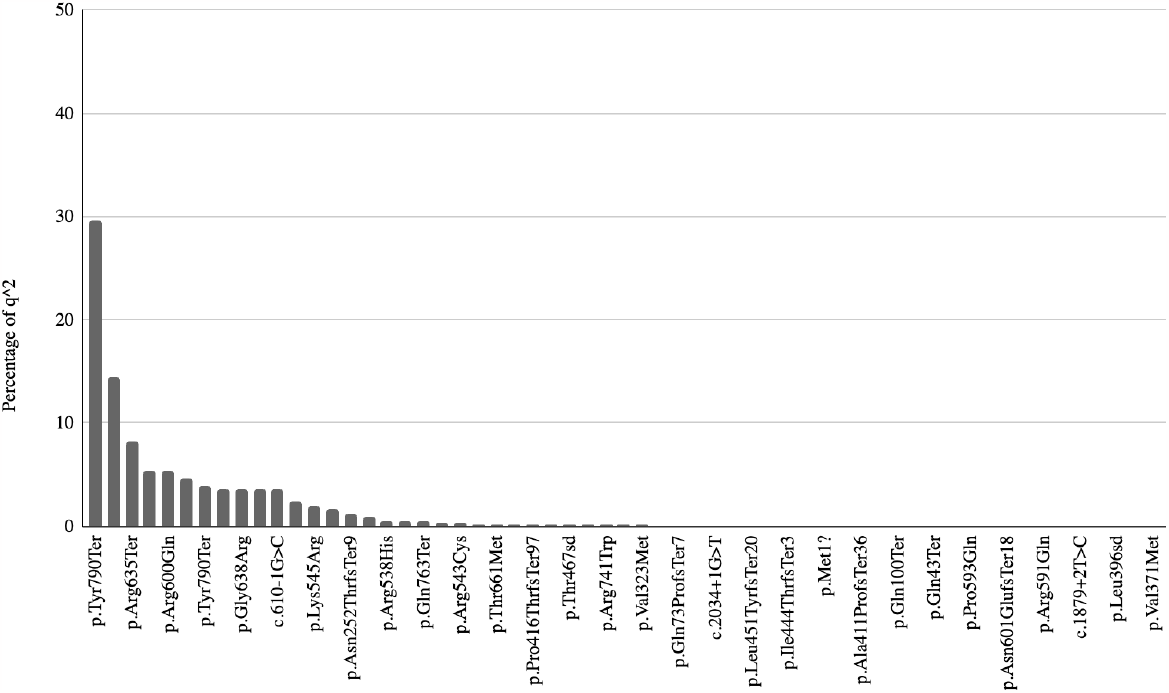
Percentage of genetic prevalence (q^2^) of pathogenic variants in PLA2G6 with non-zero allele frequencies

The most common pathogenic variant, p.Tyr790*, accounted for 11% of all non-zero allele frequency pathogenic variants; it is a truncating variant that has been identified by both ClinVar and Mastermind databases. The second most common variant, p.Val421AlafsTer26, with 7.7% allelic frequency, is a frameshift mutation leading to a truncated protein, which was an unpublished variant found in ClinVar only. The third most common variant, p.Arg635*, with 5.8% allele frequency, is also a truncated variant found in both literature and ClinVar. The fourth and fifth most common variants, p.Arg635Gln and p.Arg600Gln, are both found at 4.7% allele frequency and are missense variants located in the Patatin-like phospholipase domain.

Previously it has been shown that disease prevalence of rare diseases is directly related to the number of publications (27). Hence, we plotted the allelic frequencies of PLA2G6 pathogenic variants with the number of articles in Mastermind (Figure 3). We found a positive correlation between the number of publications and PLA2G6 allele frequencies. The seven most common pathogenic variants are labeled, showing a positive correlation between allele frequency and the number of articles published.

**Figure 3.**
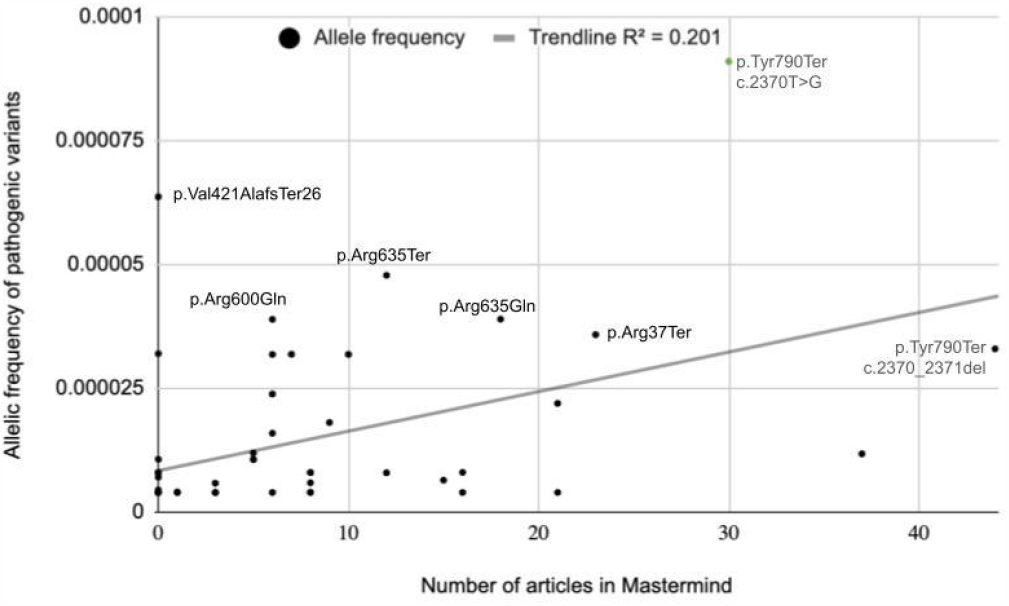
Relationship between allele frequency and the number of articles associated with the variant in Mastermind

The estimated heterozygous carrier frequency of PLA2G6 variants was found to vary between specific populations in gnomAD, as shown in Fig. 4, with the African/African American population having a significantly higher carrier frequency than other populations. The lowest carrier frequency was observed in Ashkenazi Jewish and European populations.

**Figure 4.**
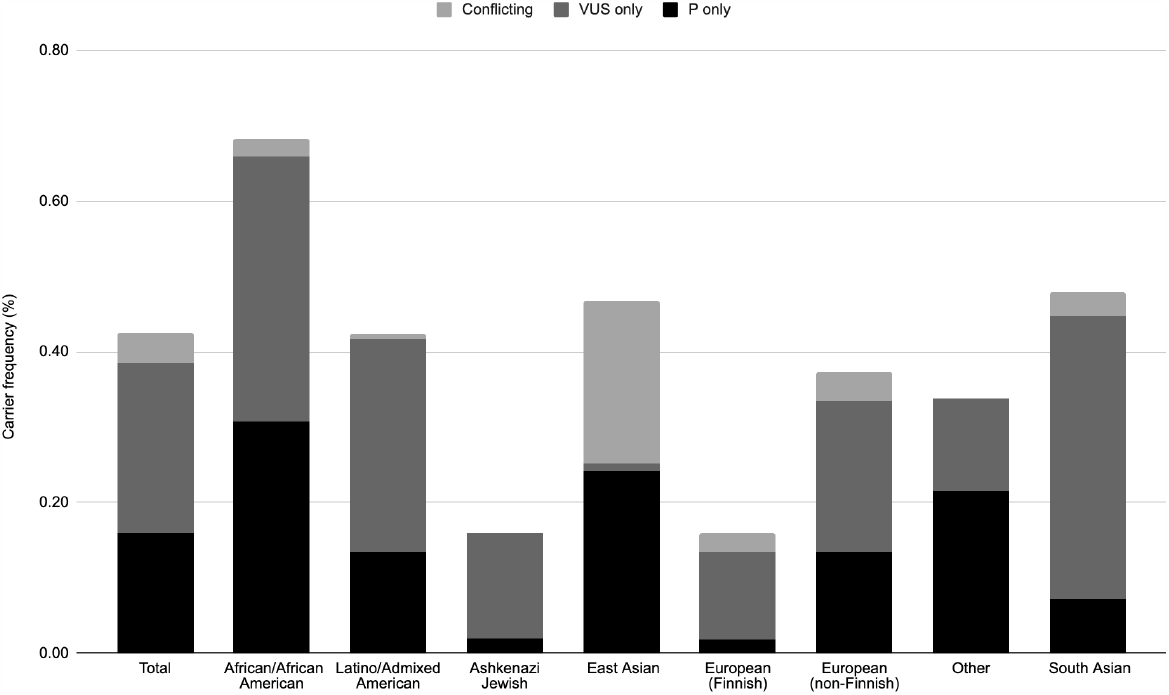
Population-Specific Carrier Frequencies of *PLA2G6* Variants. The carrier frequency was calculated using the Hardy–Weinberg equilibrium equation and the sum of the allele frequencies of the specified variants. Variants that had conflicting interpretations of pathogenicity (P vs VUS) between database and literature approach are also included in the chart.

The overall genetic prevalence of pregnancies with recessive pathogenic variants was estimated to be 6.4 pregnancies per 100,000, with the highest genetic prevalence in African populations at 23.7 pregnancies per 100,000 (Table 1). As shown in Table 1, the number of VUS variants varies significantly among populations, especially between East Asian and South Asian populations. If there are 128 million births per year worldwide, 82 (pathogenic only) to 581 (pathogenic and VUS) births with PLA2G6-associated neurodegeneration are expected annually. If we estimate the world population to be 8 billion, then the conservative estimate is that 5,210 individuals are expected to have PLA2G6-associated neurodegeneration worldwide, not accounting for the shortened life expectancy of the INAD phenotype.

**Table 1.** Carrier frequencies (%), disease prevalence per million, and genetic prevalence per million for different populations based on allelic frequencies derived from gnomAD.

|  |  | Total<br>AF | African<br>/African<br>American | Latino/<br>Admixed<br>American | Ashken<br>azi<br>Jewish | East<br>Asian | Europe<br>an<br>(Finnish) | Europe<br>an<br>(non-Finnish) | Other | South<br>Asian | Total<br>1 in |
| --- | --- | --- | --- | --- | --- | --- | --- | --- | --- | --- | --- |
| Carrier<br>frequency (%) | P+VUS+Conflicting | 0.43 | 0.68 | 0.42 | 0.16 | 0.47 | 0.16 | 0.37 | 0.34 | 0.48 | 235 |
|  | P only | 0.16 | 0.31 | 0.13 | 0.02 | 0.24 | 0.02 | 0.13 | 0.22 | 0.07 | 627 |
|  | VUS only | 0.22 | 0.35 | 0.28 | 0.14 | 0.01 | 0.12 | 0.20 | 0.12 | 0.37 | 445 |
| Genetic<br>prevalence/million | P+VUS+Conflicting | 4.54 | 11.63 | 4.48 | 0.63 | 5.47 | 0.63 | 3.49 | 2.86 | 5.73 | 220,322 |
|  | P only | 0.64 | 2.37 | 0.45 | 0.01 | 1.46 | 0.01 | 0.45 | 1.16 | 0.13 | 1,570,079 |
|  | VUS only | 1.26 | 3.08 | 2.00 | 0.49 | 0.00 | 0.33 | 1.00 | 0.38 | 3.50 | 791,498 |
| Disease<br>prevalence/million | P+VUS+Conflicting | 0.11 | 0.77 | 0.56 | 0.27 | 0.87 | 0.18 | 0.14 | 0.29 | 0.92 | 9,012,045 |
|  | P only | 0.03 | 0.21 | 0.08 | 0.01 | 0.18 | 0.00 | 0.04 | 0.20 | 0.02 | 36,542,814 |
|  | VUS only | 0.08 | 0.56 | 0.48 | 0.26 | 0.00 | 0.17 | 0.10 | 0.09 | 0.89 | 12,743,156 |

\* All variants including conflicting interpretations between two approaches (VUS vs P)

## Discussion

INAD is a rare autosomal recessive disorder caused by homozygous or compound heterozygous variants in phospholipase A2 group VI gene (*PLA2G6*) gene. Disorders associated with pathogenic variants in the PLA2G6 gene are: INAD, atypical NAD (aNAD) and PLA2G6-related dystonia-parkinsonism, also called Parkinson disease 14 (PARK14) (1). INAD patients have early-onset progressive symptoms including severe symptoms including ataxia, mental and motor deterioration, hypotonia, progressive spastic tetraparesis, visual impairments, and bulbar dysfunction. aNAD and PARK14 have a later onset of symptoms, including progressive dystonia, parkinsonism and cerebellar atrophy is a characteristic symptom (2-4). Since INAD is a rare disease, prevalence calculations have not been conducted previously. The current estimates of 1 in 1,000,000 to 1 in 2,000,000 pregnancies emphasize the urgent need for improved detection and care for individuals affected by this devastating condition. Interestingly, we found a correlation between the allele frequency and the number of literature citations, as has been previously suggested (31).

Our approach allowed us to identify variants and gather supporting evidence, leading to an improved interpretation of these variants in line with current clinical guidelines. By providing a more accurate genetic prevalence estimate through our comprehensive approach, which involves leveraging advanced tools and databases, we contribute valuable insights into the understanding and management of PLAN. Based on our analysis, we determined that the carrier frequency for PLA2G6 variants in the general population is approximately from 1 in 235 to 1 in 625 individuals, corresponding to a genetic prevalence of from 1 in 220,322 to 1 in 1,570,079 pregnancies. This estimate highlights the underdiagnosis of PLA2G6-associated neurodegeneration and underscores the need for increased awareness and diagnostic efforts. Furthermore, our study revealed a higher carrier frequency of PLA2G6 variants in African and Asian populations, stressing the importance of expanded genetic sequencing in non-European populations to ensure accurate and comprehensive diagnosis. Future research should focus on confirming our findings and implementing expanded sequencing strategies to facilitate maximal and accurate diagnosis, particularly in non-European populations.

## Data Availability

All data produced in the present work are contained in the manuscript

## Declarations

### Ethics approval and consent to participate

Not applicable.

### Consent for publication

All authors have consented to the final version of the manuscript.

### Availability of data and materials

All data analyzed during this study are available in gnomAD (https://gnomad.broadinstitute.org/) and this published article/supplementary files.

### Authors’ contributions

AK-K has analyzed the data and written the manuscript;M.S-B has extracted and analyzed data, JW extracted and analyzed data, EE has extracted and analyzed data, SMH has analyzed data and written the manuscript, LP and AH have analyzed data, MJK designed the study, has analyzed the data and written the manuscript.

## Acknowledgements

We would like to thank INADcure Foundation for helpful discussions.

## References

1. Gregory A, Kurian MA, Maher ER, et al. PLA2G6-Associated Neurodegeneration. 2008 Jun 19 [Updated 2017 Mar 23]. In: Adam MP, Feldman J, Mirzaa GM, et al., editors. GeneReviews® [Internet]. Seattle (WA): University of Washington, Seattle; 1993-2023. Available from: https://www.ncbi.nlm.nih.gov/books/NBK1675/

2. Kurian MA, Morgan N V, MacPherson L, Foster K, Peake D, Gupta R, et al. Phenotypic spectrum of neurodegeneration associated with mutations in the PLA2G6 gene (PLAN). Neurology. 2008;70(18):1623–9.

3. Iodice A, Spagnoli C, Salerno GG, Frattini D, Bertani G, Bergonzini P, et al. Infantile neuroaxonal dystrophy and PLA2G6-associated neurodegeneration: An update for the diagnosis. Brain Dev. 2017;39(2):93–100.

4. Morgan N V, Westaway SK, Morton JE V, Gregory A, Gissen P, Sonek S, et al. PLA2G6, encoding a phospholipase A2, is mutated in neurodegenerative disorders with high brain iron. Nat Genet. 2006;38(7):752–4.

5. Khateeb S, Flusser H, Ofir R, Shelef I, Narkis G, Vardi G, et al. PLA2G6 mutation underlies infantile neuroaxonal dystrophy. Am J Hum Genet. 2006;79(5):942–8.

6. Paisan-Ruiz C, Bhatia KP, Li A, Hernandez D, Davis M, Wood NW, et al. Characterization of PLA2G6 as a locus for dystonia-parkinsonism. Ann Neurol. 2009;65(1):19–23.

7. Sumi-Akamaru H, Beck G, Kato S, Mochizuki H. Neuroaxonal dystrophy in PLA2G6 knockout mice. Neuropathology. 2015;35(3):289–302.

8. Riku Y, Ikeuchi T, Yoshino H, Mimuro M, Mano K, Goto Y, et al. Extensive aggregation of α-synuclein and tau in juvenile-onset neuroaxonal dystrophy: an autopsied individual with a novel mutation in the PLA2G6 gene-splicing site. Acta Neuropathol Commun. 2013;1:12.

9. Paisán-Ruiz C, Li A, Schneider SA, Holton JL, Johnson R, Kidd D, et al. Widespread Lewy body and tau accumulation in childhood and adult onset dystonia-parkinsonism cases with PLA2G6 mutations. Neurobiol Aging. 2012;33(4):814–23.

10. Babin PL, Rao SNR, Chacko A, Alvina FB, Panwala A, Panwala L, et al. Infantile Neuroaxonal Dystrophy: Diagnosis and Possible Treatments. Front Genet. 2018;9.

11. Lin G, Tepe B, McGrane G, Tipon RC, Croft G, Panwala L, et al. Exploring therapeutic strategies for infantile neuronal axonal dystrophy (INAD/PARK14). Elife. 2023;12:e82555.

12. Vinkšel M, Writzl K, Maver A, Peterlin B. Improving diagnostics of rare genetic diseases with NGS approaches. J Community Genet. 2021;12(2):247–256

13. Krude H, Mundlos S, Øien NC, Opitz R, Schuelke M. What can go wrong in the non-coding genome and how to interpret whole genome sequencing data. Medizinische Genetik. 2021;14:2071.

14. Landrum MJ, Chitipiralla S, Brown GR, Chen C, Gu B, Hart J, et al. ClinVar: improvements to accessing data. Nucleic Acids Res. 2020 Jan 8;48(D1):D835–44.

15. Singer-Berk M, Gudmundsson S, Baxter S, Seaby EG, England E, Wood JC, Son RG, Watts NA, Karczewski KJ, Harrison SM, MacArthur DG, Rehm HL, O’Donnell-Luria A. Advanced variant classi?cation framework reduces the false positive rate of predicted loss-of-function variants in population sequencing data. Am J Hum Genet. 2023 Sep 7;110(9):1496–1508.

16. Smith CIE, Bergman P, Hagey DW. Estimating the number of diseases - the concept of rare, ultra-rare, and hyper-rare. iScience. 2022;25(8):104698.

17. Infantile Neuroaxonal Dystrophy (INAD), Accessed on 11/10/2023: https://my.clevelandclinic.org/health/diseases/22767-infantile-neuroaxonal-dystrophy-inad

18. Xiao Q, Lauschke VM. The prevalence, genetic complexity and population-specific founder effects of human autosomal recessive disorders. NPJ Genom Med. 2021;6(1):41.

19. Chunn LM, Nefcy DC, Scouten RW, Tarpey RP, Chauhan G, Lim MS, et al. Mastermind: A Comprehensive Genomic Association Search Engine for Empirical Evidence Curation and Genetic Variant Interpretation. Front Genet. 2020;11.

20. Xiao Q, Lauschke VM. The prevalence, genetic complexity and population-specific founder effects of human autosomal recessive disorders. NPJ Genom Med. 2021;6(1):41.

21. den Dunnen JT, Dalgleish R, Maglott DR, Hart RK, Greenblatt MS, McGowan-Jordan J, et al. HGVS Recommendations for the Description of Sequence Variants: 2016 Update. Hum Mutat. 2016;37(6):564–9.

22. Richards S, Aziz N, Bale S, Bick D, Das S, Gastier-Foster J, et al. Standards and guidelines for the interpretation of sequence variants: a joint consensus recommendation of the American College of Medical Genetics and Genomics and the Association for Molecular Pathology. Genet Med. 2015;17(5):405–24.

23. Karczewski KJ, Francioli LC, Tiao G, Cummings BB, Alföldi J, Wang Q, et al. The mutational constraint spectrum quantified from variation in 141,456 humans. Nature. 2020;581(7809):434–43.

24. Chunn LM, Bissonnette J, Heinrich S V, Mercurio SA, Kiel MJ, Rutsch F, et al. Estimation of ENPP1 deficiency genetic prevalence using a comprehensive literature review and population databases. Orphanet J Rare Dis. 2022;17(1):421.

25. Fan S, Zhao T, Sun L. The global prevalence and ethnic heterogeneity of iron-refractory iron deficiency anaemia. Orphanet J Rare Dis 2023;18:2.

26. Hardy GH. Mendelian Proportions in a Mixed Population. Science 1979;28(706):49–50.

27. Sim NL, Kumar P, Hu J, Henikoff S, Schneider G, Ng PC. SIFT web server: predicting effects of amino acid substitutions on proteins. Nucleic Acids Res. 2012;40(W1):W452–7.

28. Schwarz JM, Cooper DN, Schuelke M, Seelow D. MutationTaster2: mutation prediction for the deep-sequencing age. Nat Methods. 2014;11(4):361–2.

29. Jian X, Boerwinkle E, Liu X. In silico prediction of splice-altering single nucleotide variants in the human genome. Nucleic Acids Res. 2014;42(22):13534–44.

30. Adzhubei I, Jordan DM, Sunyaev SR. Predicting Functional Effect of Human Missense Mutations Using PolyPhen-2. Curr Protoc Hum Genet. 2013;76(1).

31. Shourick, J., Wack, M. & Jannot, AS. Assessing rare diseases prevalence using literature quantification. Orphanet J Rare Dis 2021;16:139.

